# The Coronavirus Health and Impact Survey (CRISIS) reveals reproducible correlates of pandemic-related mood states across the Atlantic

**DOI:** 10.1101/2020.08.24.20181123

**Authors:** Aki Nikolaidis, Diana Paksarian, Lindsay Alexander, Jacob Derosa, Julia Dunn, Dylan M. Nielson, Irene Droney, Minji Kang, Ioanna Douka, Evelyn Bromet, Michael Milham, Argyris Stringaris, Kathleen R. Merikangas

**Affiliations:** Center for the Developing Brain, The Child Mind Institute, New York, NY; Genetic Epidemiology Research Branch, Intramural Research Program, National Institute of Mental Health, Bethesda, MD; Section on Clinical and Computational Psychiatry (Compψ), National Institute of Mental Health, National Institutes of Health, Bethesda, Maryland; Department of Psychiatry, Renaissance School of Medicine at Stony Brook University; Nathan Kline Institute for Psychiatric Research; Johns Hopkins Bloomberg School of Public Health

## Abstract

The COVID-19 pandemic and its social and economic consequences have had adverse impacts on physical and mental health worldwide and exposed all segments of the population to protracted uncertainty and daily disruptions. The CoRonavIruS health and Impact Survey (CRISIS) was developed for use as an easy to implement and robust questionnaire covering key domains relevant to mental distress and resilience during the pandemic. In the current work, we demonstrate the feasibility, psychometric structure and construct validity of this survey. We then show that pre-existing mood states, perceived COVID risk, and lifestyle changes are strongly associated with negative mood states during the pandemic in population samples of adults and in parents reporting on their children in the US and UK. Ongoing studies using CRISIS include international studies of COVID-related ill health conducted during different phases of the pandemic and follow-up studies of cohorts characterized before the COVID pandemic.

## Introduction

Since its first documented occurrence in December 2019, COVID-19 has taken an enormous toll on human life and health and is in line to become the leading cause of death in many countries, including the US. Along with the immediate health impacts, the virus and prevention strategies have perturbed the core structure of daily life, including financial security, work, school, recreation, and social interactions. The COVID-19 pandemic stands in stark contrast to recent epidemics such as SARS and MERS in terms of the total number of cases and deaths^1^. Prior mass disasters (World Trade Center attacks, mass shootings), natural disasters (hurricanes, floods) and environmental exposures (oil spills, radiation exposures) have been associated with increases in depression, posttraumatic stress disorder (PTSD), substance use, generalized anxiety disorder and a range of other mental health outcomes.^2-6^ These studies were conducted primarily in the aftermath of these catastrophes. Much less is known about the risk and protective factors for well-being during and after prolonged threats^3^, like the COVID-19 pandemic, which continues to unfold.

The pernicious mental health effects of the COVID-19 pandemic may result from death of loved ones, disease severity, social isolation and quarantine, unemployment, financial hardship, domestic violence, and educational disruptions. Each of these factors is independently associated with psychological comorbidities.^7-13^ Apart from studies of the neuropsychiatric impact of COVID-19 on health care workers and people who contracted the virus,^9,14,15^ there are a growing number of international longitudinal surveys designed to document community-level pandemic-related psychological distress. Many of these surveys have been adapted to encompass a wider range of mental health outcomes than measured in previous epidemics^4^, with the most common domains including stress, anxiety, loneliness, depression, social support, media and technology use, sleep, and post-traumatic stress. Most of these surveys included established symptom scales, such as the Generalized Anxiety Disorder (GAD-7), Patient Health Questionnaire (PHQ-9), and UCLA Loneliness Scale,^14,16,17^ as documented by the COVID-MINDS network of longitudinal studies on the global mental health impact of COVID-19). Published findings from these studies have shown high levels of anxiety and depression symptoms post COVID-19 based on cut-points from US and European sources. However, the most robust risk factors for disaster-related mental ill health – prior psychopathology and exposure severity^26^ – remain largely understudied to date. With some exceptions^18-24^ few of the COVID-19 specific assessment tools developed to track mental health responses to the COVID-19 pandemic have been psychometrically validated.

Aligning with NIH COVID-19 research priorities,^25,26^ the Coronavirus Health and Impact Survey (CRISIS) Initiative was established as a collaborative and multidisciplinary effort to identify predictors of acute and long-term psychopathology. Key predictors include impairment and disability associated with the COVID-19 pandemic in samples that were well-characterized prior to the pandemic across the globe. The first step was to develop, pilot, and test the psychometric properties of a comprehensive instrument that captured a core set of domains. Specifically, the CRISIS instrument was designed to assess pertinent mental, behavioral, and physical health domains that capture the multi-level emotional and behavioral impact of the pandemic, as well as a range of pandemic-related and pre-existing risk and protective factors. The CRISIS includes forms for adults ages 19-64, parent reports for children aged 9-18, and youth aged 9-18. The following domains are assessed: (1) background and demographic characteristics, including household composition and crowding; (2) physical and mental health 3 months prior to the pandemic; (3) COVID-19 exposure and infection status; (4) life changes due to the pandemic; (5) concerns and worries associated with COVID-19; (6) current well-being determined by the circumplex model of affect;^27,28^ and (7) behavioral factors, such as media use, sleep, physical activity, and substance use. We also developed a short form of the CRISIS for follow up of the samples that excludes the background and the 3-months prior physical and mental health sections. Both the baseline and follow up surveys are licensed on Creative Commons (CC) BY4.0 and are available for download at crisissurvey.org.

This article describes the properties of the CRISIS in relatively large (n = 5,646) pilot samples of adults and parents in the United States (US) and United Kingdom (UK) collected in April 2020. Across the multiple samples, the aims were to: 1) describe the CRISIS and assess its acceptability and feasibility, 2) evaluate the factor structure of the major domains; 3) examine the test-retest reliability and construct validity of these domains across the multiple samples, and 4) estimate the relative importance of the measured domains to current mood states (operationalized as mood over the previous 2 weeks).

## 2 Methods

### 2.1 Samples

Pilot data were collected between April 7th and 17th, 2020, through Prolific Academic (https://www.prolific.ac/) (PA), an online crowdsourced survey recruitment service. Participants who signed up to join the PA participant pool received monetary compensation for their time. PA participants have been shown to be more diverse and provide higher quality data than similar data collection platforms.^29^ We requested 3,000 adult self-report and 3,000 parent-report participants in the US and the UK. For parent reports, users were screened based on having a child between 5 and 17 years old, and reported on their oldest child in that age range. Portions of the sample were targeted at regions that were more severely impacted by COVID-19 in late March 2020 (New York, California, London, and Manchester).

We received a total of 5,928 unduplicated responses, from which we dropped 282 with incomplete forms. The final analytic sample sizes were 1,527 US adults (231 California; 246 New York), 1,539 UK adults (248 London; 238 Manchester), 1,121 US parents (27 California; 19 New York), and 1,459 UK parents (172 London; 219 Manchester). Samples were further divided into training (⅔) and hold-out (⅓) samples for assessing the reproducibility of associations with current mood states. Resulting training data sample sizes were 935 (US Adult), 938 (UK adult), 673 (US Parent), and 877 (UK Parent). Separate 24-hour test-retest reliability samples were obtained from 74 US adults, 76 UK adults, 71 US parents, and 75 UK parents concurrently with the main US/UK samples.

Because all data were collected anonymously, no IRB oversight was required. Exemption from IRB oversight was approved by the Advarra Institutional Review Board. Participants using the PA website are required to agree to the Terms of Service notification (https://prolific.ac/assets/docs/Participant_Terms.pdf) before being allowed to complete surveys. Per the IRB exemption, no additional informed consent was required.

### 2.2. Measurement domains

To assess the structure, psychometric properties, and construct validity of the CRISIS, we focused on the following domains and indicators (see Supplement for more details):

#### Participant characteristics

Age, sex, race/ethnicity, self- or parent-rated health, urbanicity, education, household size, health insurance coverage, and family’s receipt of government assistance. Race was reported to Prolific Academic and combined with a question on Hispanic ethnicity to generate the following categories: Hispanic, non-Hispanic white, non-Hispanic black, Asian, and other.

#### SARS-CoV-2 exposure/infection in the past 2 weeks

Possible exposure to SARS-CoV-2, possible symptoms of COVID-19, family member diagnosis of COVID-19, essential worker in the household, and whether there had been any impacts on family members such as hospitalization, quarantine, and job loss because of COVID-19.

#### COVID Worries in the past 2 weeks

Participants reported on a 5-point Likert scale how worried they have been during the past 2 weeks about infection, friends and family being infected, and possible impacts on physical and mental health, as well as time spent reading or talking about COVID-19, and hope that the pandemic will end soon.

#### Life Changes due to the pandemic in the past 2 weeks

Downstream and subjective impacts of structural changes, such as changes in social contacts, effects on family relationships, changes in living situation, food insecurity, and stressors associated with these changes (14 items). Participants were also asked about job loss and school closure due to the pandemic; these items were used as internal validators.

#### Mood States

Ten items from the circumplex model of affect^27,28^ were included to measure mood/anxiety, both during the past 2 weeks (hereafter referred to as “Current Mood States”) and during the 3 months prior to the pandemic (hereafter referred to as “Prior Mood States”).

#### Substance Use

Frequency of use of tobacco, alcohol, marijuana, and other substances during the past 2 weeks and during the 3 months prior to the pandemic.

#### Daily Behaviors

Average weekday and weekend bedtime and sleep duration, frequency of exercise, time spent outdoors, and length of media use per day were rated for the past 2 weeks and the 3 months prior to the pandemic.

### 2.3. Analysis

#### Overview

Analyses focused on 5 domains of interest: COVID Worries, Life Changes, Mood States, Substance Use, and Daily Behaviors. The statistical approaches are described below, with additional details in the supplement. Structure was assessed via factor analysis and community detection subtyping. Test-retest reliability was measured via the Intraclass Correlation Coefficient (ICC(3,1)).^30^ Construct validity was assessed by comparing associations between domains using chi-squared tests, ANOVAs and ANCOVAs, and via random forests.

#### Factor Analysis (Aim 2)

Confirmatory factor analysis (CFA) was performed in each sample. To assess the stability and reproducibility of our factor structure, we split the ⅔ training dataset further into two datasets each corresponding to ⅓ of the full dataset, we performed CFA on each. To assess unidimensionality, CFA was applied in each sample split, with a comparative fit index (CFI) of > 0.95 and an Omega of > 0.8 indicating adequate fit.^31^ Resulting factor scores were used to assess construct validity for Aim 4.

#### Community Detection Based Subtyping (Aim 2)

Louvain community detection (LCD)^32^ was used to derive data-driven subtypes on domains that exhibited poor unidimensional fit in CFA. In order to maximize the modularity of the sample, LCD selects the cluster resolution that maximizes the within-community coherence and between-community segregation. LCD^32^ was enhanced through bootstrap aggregation (i.e., bagging) which has been shown to generate more reproducible clusters (see Supplement).^33,34^ Resulting subtypes were used to assess construct validity for Aim 4.

#### Test-Retest Reliability (Aim 3)

We assessed the reliability of the factor scores and individual items using intraclass correlation coefficient (ICC 3,1)^30^ on the separate 24 hour test retest sample for each of the US and UK adult and parent report samples. ICC results for factors and individual items are summarized in the Supplement.

#### Random Forests (Aim 4)

Random forest (RF), a robust technique known for its ability to model dependencies between predictor variables (See Supplement),^35,36^ was used to examine associations of participant characteristics with Mood States and Behaviors (e.g., COVID Worries, Life Changes, Daily Behaviors, Media Use, Substance Use, and Prior Mood States). RF assesses performance across the ensemble of decision trees on the samples not included in each bootstrap iteration. The out-of-bag mean square error (MSE) and node impurity were used as measures of relative variable importance for each predictor. Generalizability of the importance of the variables identified in each random forest analysis was assessed in the ⅓ hold-out datasets. In each subsample, a linear regression model was trained using the four most important variables and their interaction terms. These models were then applied to each corresponding hold-out set to evaluate out-of-sample performance.

## 3. Results

### 3.1 Acceptability and Feasibility (Aim 1)

Characteristics of the total analytic sample are presented in Table 1. Supplemental Table 1 shows the amount of missing data and time to completion. The proportion of complete surveys was high (95.2%). The numbers of missing items per survey were low, on average 0.6 (SD = 1.8) and 0.5 (SD = 1.1) for the US and UK Adult reports respectively, and 0.4 (SD = 0.9) and 0.4 (SD = 1.0) for the US and UK parent reports. There was an average of 13.9 (SD = 13.1) and 14.4 (SD = 11.5) minutes to completion for the US and UK Adult reports respectively and 14.1 (SD = 7.5) and 13.9 (SD = 20.9) for the US and UK parent reports. Feedback from the open-ended questions was generally positive, and no comments suggested that CRISIS was a burden, consistent with the high completion rate and low rate of missing values.

**Table 1:**
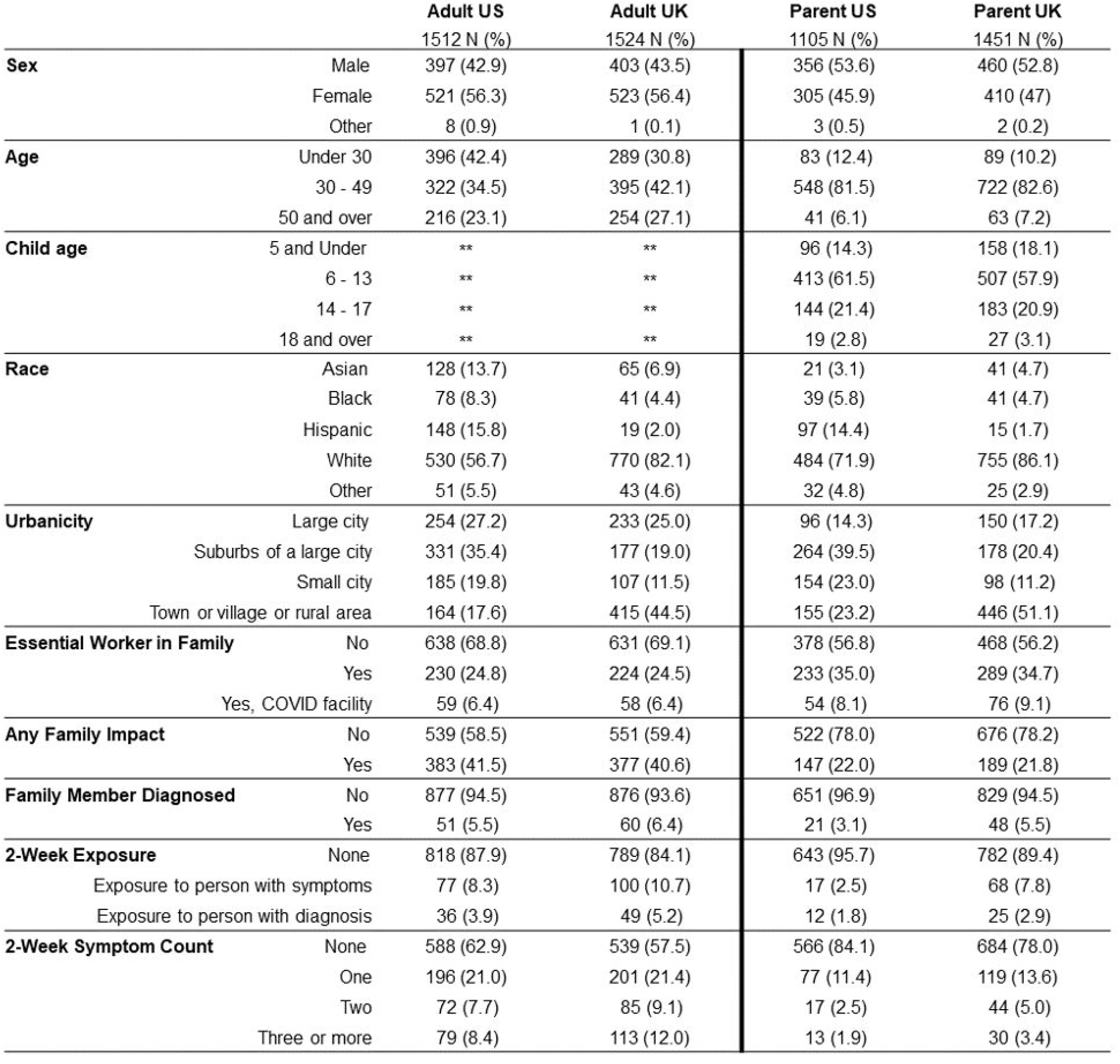
Frequencies and percentages of key demographic variables and COVID-related experiences by sample.

|  |  | Adult US<br>1512 N (%) | Adult UK<br>1524 N (%) | Parent US<br>1105 N (%) | Parent UK<br>1451 N (%) |
| --- | --- | --- | --- | --- | --- |
| **Sex** | Male | 397 (42.9) | 403 (43.5) | 356 (53.6) | 460 (52.8) |
|  | Female | 521 (56.3) | 523 (56.4) | 305 (45.9) | 410 (47) |
|  | Other | 8 (0.9) | 1 (0.1) | 3 (0.5) | 2 (0.2) |
| **Age** | Under 30 | 396 (42.4) | 289 (30.8) | 83 (12.4) | 89 (10.2) |
|  | 30 - 49 | 322 (34.5) | 395 (42.1) | 548 (81.5) | 722 (82.6) |
|  | 50 and over | 216 (23.1) | 254 (27.1) | 41 (6.1) | 63 (7.2) |
| **Child age** | 5 and Under | ** | ** | 96 (14.3) | 158 (18.1) |
|  | 6 - 13 | ** | ** | 413 (61.5) | 507 (57.9) |
|  | 14 - 17 | ** | ** | 144 (21.4) | 183 (20.9) |
|  | 18 and over | ** | ** | 19 (2.8) | 27 (3.1) |
| **Race** | Asian | 128 (13.7) | 65 (6.9) | 21 (3.1) | 41 (4.7) |
|  | Black | 78 (8.3) | 41 (4.4) | 39 (5.8) | 41 (4.7) |
|  | Hispanic | 148 (15.8) | 19 (2.0) | 97 (14.4) | 15 (1.7) |
|  | White | 530 (56.7) | 770 (82.1) | 484 (71.9) | 755 (86.1) |
|  | Other | 51 (5.5) | 43 (4.6) | 32 (4.8) | 25 (2.9) |
| **Urbanicity** | Large city | 254 (27.2) | 233 (25.0) | 96 (14.3) | 150 (17.2) |
|  | Suburbs of a large city | 331 (35.4) | 177 (19.0) | 264 (39.5) | 178 (20.4) |
|  | Small city | 185 (19.8) | 107 (11.5) | 154 (23.0) | 98 (11.2) |
|  | Town or village or rural area | 164 (17.6) | 415 (44.5) | 155 (23.2) | 446 (51.1) |
| **Essential Worker in Family** | No | 638 (68.8) | 631 (69.1) | 378 (56.8) | 468 (56.2) |
|  | Yes | 230 (24.8) | 224 (24.5) | 233 (35.0) | 289 (34.7) |
|  | Yes, COVID facility | 59 (6.4) | 58 (6.4) | 54 (8.1) | 76 (9.1) |
| **Any Family Impact** | No | 539 (58.5) | 551 (59.4) | 522 (78.0) | 676 (78.2) |
|  | Yes | 383 (41.5) | 377 (40.6) | 147 (22.0) | 189 (21.8) |
| **Family Member Diagnosed** | No | 877 (94.5) | 876 (93.6) | 651 (96.9) | 829 (94.5) |
|  | Yes | 51 (5.5) | 60 (6.4) | 21 (3.1) | 48 (5.5) |
| **2-Week Exposure** | None | 818 (87.9) | 789 (84.1) | 643 (95.7) | 782 (89.4) |
|  | Exposure to person with symptoms | 77 (8.3) | 100 (10.7) | 17 (2.5) | 68 (7.8) |
|  | Exposure to person with diagnosis | 36 (3.9) | 49 (5.2) | 12 (1.8) | 25 (2.9) |
| **2-Week Symptom Count** | None | 588 (62.9) | 539 (57.5) | 566 (84.1) | 684 (78.0) |
|  | One | 196 (21.0) | 201 (21.4) | 77 (11.4) | 119 (13.6) |
|  | Two | 72 (7.7) | 85 (9.1) | 17 (2.5) | 44 (5.0) |
|  | Three or more | 79 (8.4) | 113 (12.0) | 13 (1.9) | 30 (3.4) |

### 3.2 Factor Analysis and Community Detection Subtyping (Aim 2)

To evaluate the structure of the 5 domains of interest, we first conducted confirmatory factor analysis (Table 2). The Mood States and COVID Worries domains each demonstrated high internal consistency as assessed using coefficient Omega (>0.8), and good unidimensional model fit as assessed using the comparative fit index (CFI >0.95) across each split samples of US and UK in both the Adult Self-Report and Parent-Report data. Associations between factor scores are depicted in Supplementary Figure 2. We did not find strong evidence for unidimensional model fit for Daily Behaviors and Media Use (CFI < 0.9), Life Changes (Adult CFI < 0.9; Parent CFI <0.95), or adult Substance Use (CFI > 0.95 in US and <0.75 in UK), which was expected given that these domains were designed to capture a broad variety of behaviors.

**Table 2:**
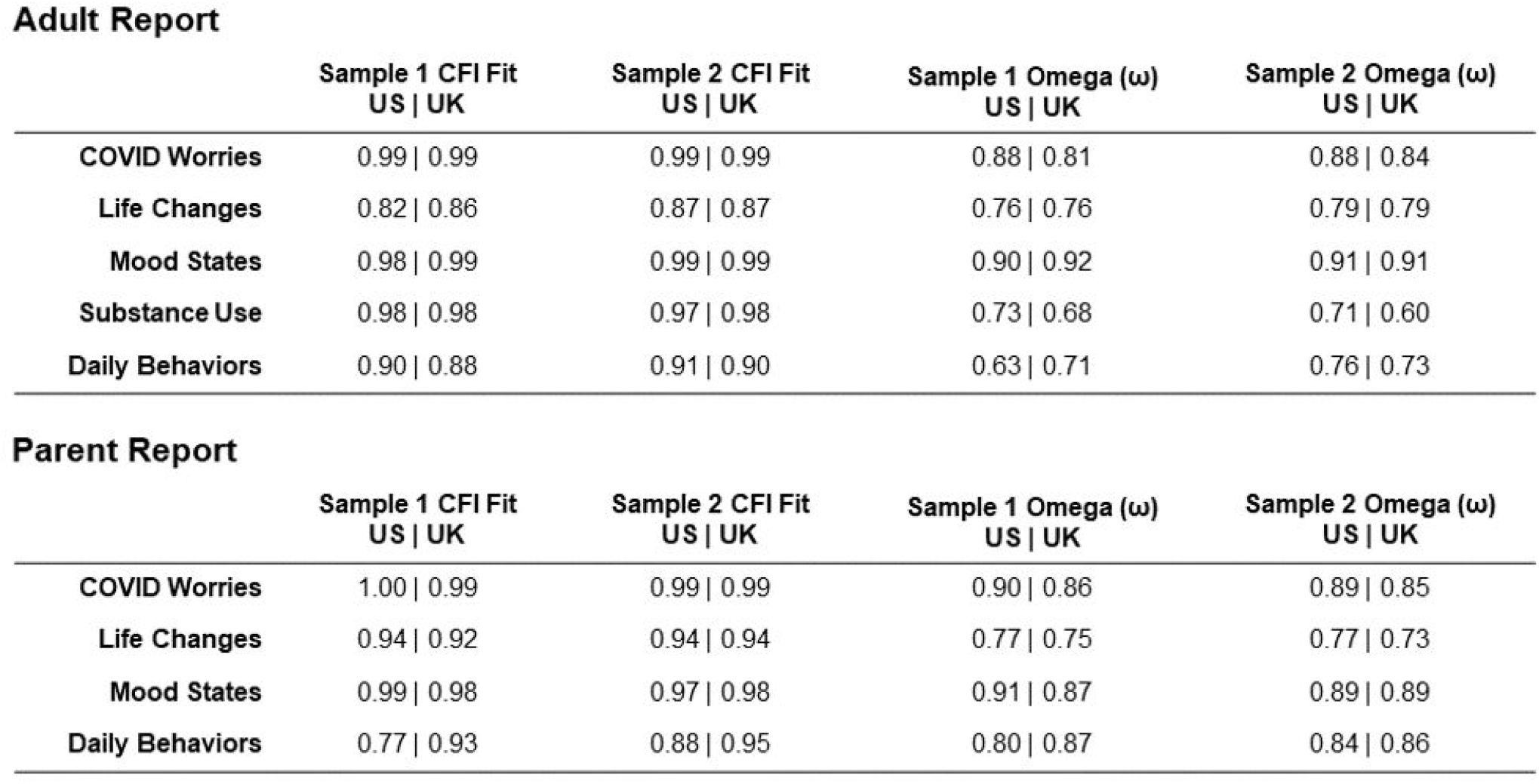
Fit statistics from confirmatory factor analysis in split-half samples from adult self-report and parent respondents in the US and UK.

Adult Report
| | Sample 1 CFI Fit<br>US \| UK | Sample 2 CFI Fit<br>US \| UK | Sample 1 Omega ($\omega$)<br>US \| UK | Sample 2 Omega ($\omega$)<br>US \| UK |
| --- | --- | --- | --- | --- |
| COVID Worries | 0.99 \| 0.99 | 0.99 \| 0.99 | 0.88 \| 0.81 | 0.88 \| 0.84 |
| Life Changes | 0.82 \| 0.86 | 0.87 \| 0.87 | 0.76 \| 0.76 | 0.79 \| 0.79 |
| Mood States | 0.98 \| 0.99 | 0.99 \| 0.99 | 0.90 \| 0.92 | 0.91 \| 0.91 |
| Substance Use | 0.98 \| 0.98 | 0.97 \| 0.98 | 0.73 \| 0.68 | 0.71 \| 0.60 |
| Daily Behaviors | 0.90 \| 0.88 | 0.91 \| 0.90 | 0.63 \| 0.71 | 0.76 \| 0.73 |

| | Sample 1 CFI Fit<br>US \| UK | Sample 2 CFI Fit<br>US \| UK | Sample 1 Omega ($\omega$)<br>US \| UK | Sample 2 Omega ($\omega$)<br>US \| UK |
| --- | --- | --- | --- | --- |
| COVID Worries | 1.00 \| 0.99 | 0.99 \| 0.99 | 0.90 \| 0.86 | 0.89 \| 0.85 |
| Life Changes | 0.94 \| 0.92 | 0.94 \| 0.94 | 0.77 \| 0.75 | 0.77 \| 0.73 |
| Mood States | 0.99 \| 0.98 | 0.97 \| 0.98 | 0.91 \| 0.87 | 0.89 \| 0.89 |
| Daily Behaviors | 0.77 \| 0.93 | 0.88 \| 0.95 | 0.80 \| 0.87 | 0.84 \| 0.86 |

Because the Life Changes, Substance Use, and Behavior & Media Use domains generally exhibited poor unidimensional fit in CFA, they were summarized via community detection subtyping. We conducted 2 subtyping analyses: one focused on the Life Changes domain, and another focused on the combined Substance Use and Behavior & Media Use domains, using questions pertaining to the 3 months prior to the pandemic (referred to as Prior Habits). The derived Life Changes subtypes in adults and children are displayed in Figure 1. For both the adult self-report and parent report, the Life Changes subtypes were highly reproducible across the US and UK samples, as Pearson’s correlations across the US and UK profiles show high consistency (r = 0.91-0.99). Prior Habits subtypes in adults and children are described in the supplement.

**Figure 1:**
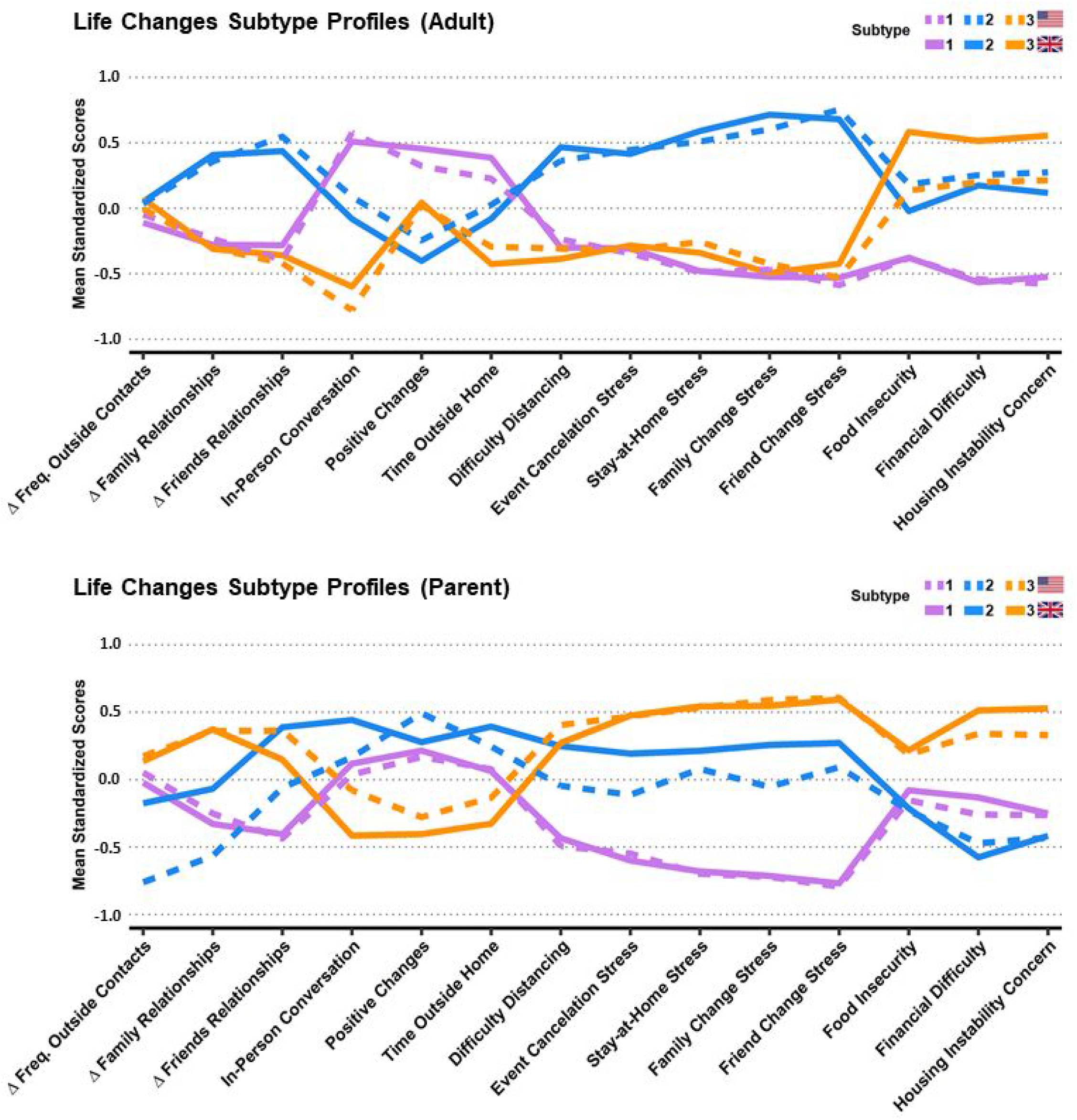
Life Changes Subtype profiles from adult self-reports and parent reports. Mean normalized profile loadings are displayed on the y-axis. US subtypes in solid lines, UK in dashed lines. Adult Subtypes: Purple (1): *low stress*, Blue (2): *social/interpersonal stress*, Orange (3): *economic stress*. Parent-Report Subtypes: Purple (1): *low stress*, Blue (2): *social stress*, Orange (3): *social/economic stress* Notes: Δ Family Relationships and Δ Friends Relationships are coded so that higher scores indicate worsening relationship quality of. Prior to the community detection analyses In-Person Conversation was re-coded into tertiles.

In the adult sample, the *low stress* subtype (purple; 1) reported greater positive changes, in-person conversations, and time outside; lower levels of stress from distancing, cancellations, and relationship changes; and lower levels of food insecurity, financial difficulty, and housing instability. The *social/interpersonal stress* subtype (blue; 2) reported worsening of relationships, higher stress levels, few positive changes, and moderate levels of economic concerns. The *economic stress* subtype (orange; 3) reported the most problems with food security, financial difficulty, and housing stability, but lower levels of other stresses, while also having the least in-person conversations.

In the parent report, the *low stress* subtype (purple; 1) had somewhat improved family and friend relationships, low stress related to social and interpersonal changes, and average levels of economic stress, time outside, and positive changes. The *social stress* subtype (blue; 2) reported moderate to high levels of stress related to social and interpersonal changes; low levels of economic stress; higher levels of positive changes, time outside, and in-person conversations; US parents reported more worsening of relationships than did UK parents. The *social/economic stress* subtype (orange; 3) reported the highest levels of economic and social/interpersonal stress, worsening of family relationships, and the lowest levels of positive changes, time outside the home, and in-person conversations.

### 3.3 Test-Retest Reliability (Aim 3)

24-hour test-retest reliability results are presented in Supplemental Table 4. We found the Mood States and COVID Worries factor reliabilities were high (ICC (3,1) = 0.79-0.87) in all Adult and Parent Report samples. Reliability of single items not included in factor scores was generally moderate to excellent (ICC of Prior Habits variables mean = 0.79, sd = 0.09; ICC of Life Changes variables mean = 0.64, sd = 0.15; ICC of Substance Use variables mean = 0.88, sd = 0.19) and are presented in Supplementary Table 2.

### 3.4 Associations Between Domains (Aim 4)

Mean factor scores by participant characteristics among adults are presented in Table 3. Most associations were consistent across US and UK adults. COVID Worries was consistently higher among those with any family impact (US p=.001; UK p <.001), a family member with a COVID diagnosis (US p=.019; UK p=.005), and potential symptoms (US p=0.001; UK p<.001). In addition to associations with age and sex, Mood States scores were also consistently higher among those with any family impact (US p=.011; UK p=.005), exposure to someone with symptoms (US p=.036; UK p=.038), and potential symptoms (US p<.001; UK p<.001). COVID Worries and Mood States factor scores were not associated with participant race/ethnicity or having an essential worker in the home.

**Table 3:**
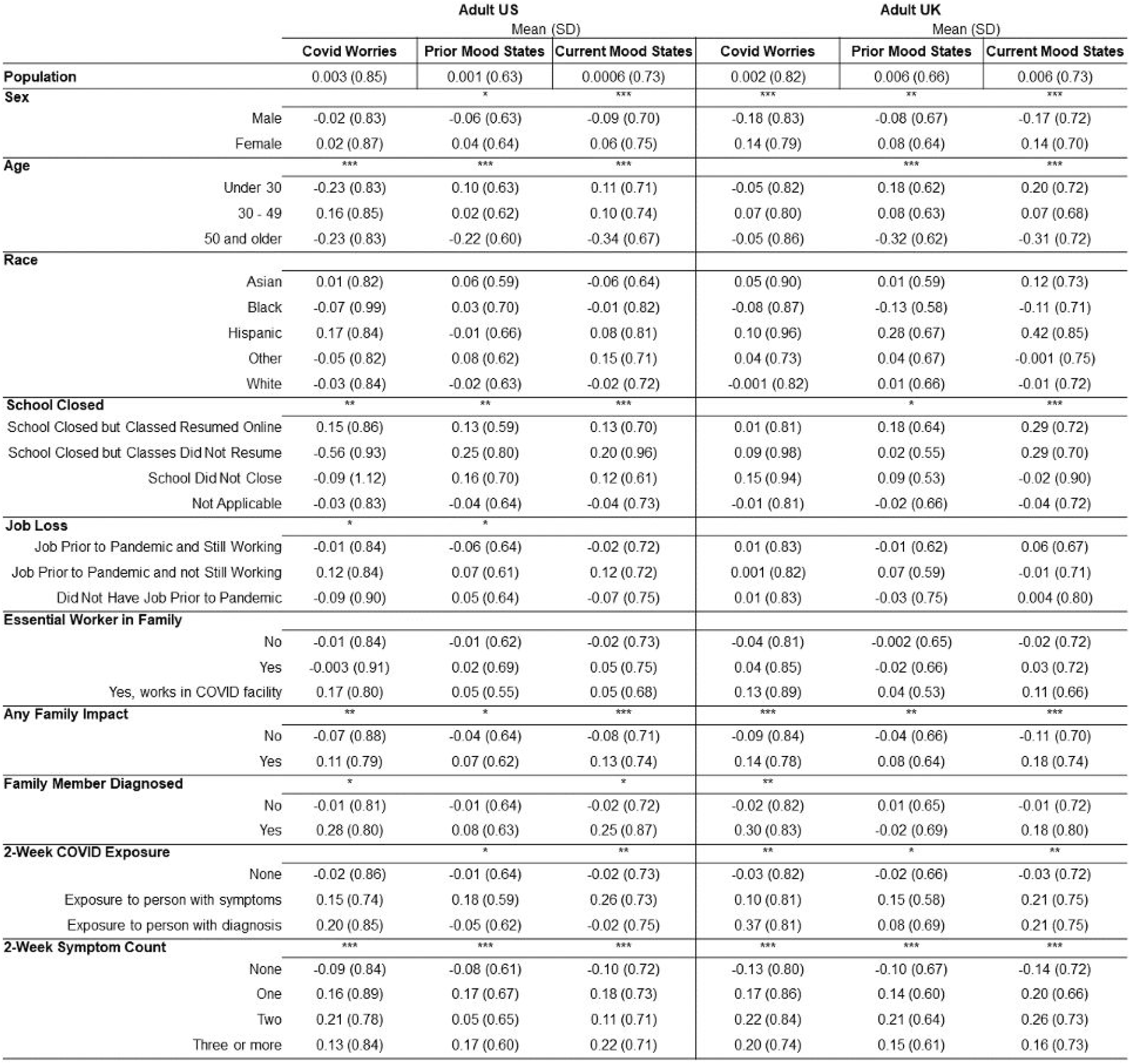
Mean factor scores for unidimensional constructs among adults by demographic and COVID-related characteristics. Significant group differences are represented by asterisks, uncorrected for multiple comparisons: * *p <*.05, ** *p* <.01, \*\*\**p* <.001.

Associations of Life Changes subtypes with factor scores, participant characteristics, and school closure and job loss are presented in Table 4. Life changes subtype was associated with Mood States (Prior: US p< 0.00001, UK p< 0.00001; Current: US p > 0.00001, UK p > 0.00001), as well as COVID Worries score (US p < 0.00001, UK p < 0.00001). Adjusting for Prior Mood States, Current Mood States scores were highest in the *social/interpersonal stress* subtype among adults and the *social/economic stress* subtype among parent reports (Table 4). Life Changes subtypes also differed by key demographic characteristics including age, race/ethnicity, education, rooms in house, household density, and employment (see Table 4). Corresponding results for Prior Habits subtypes appear in Supplemental Tables 3 and 4. Briefly, we find differences between Prior Habit subtypes in COVID Worries, Prior Mood States, and Current Mood States, indicating the importance of prior behavioral and psychological states in influencing the negative mental health outcomes of the pandemic.

**Table 4.**
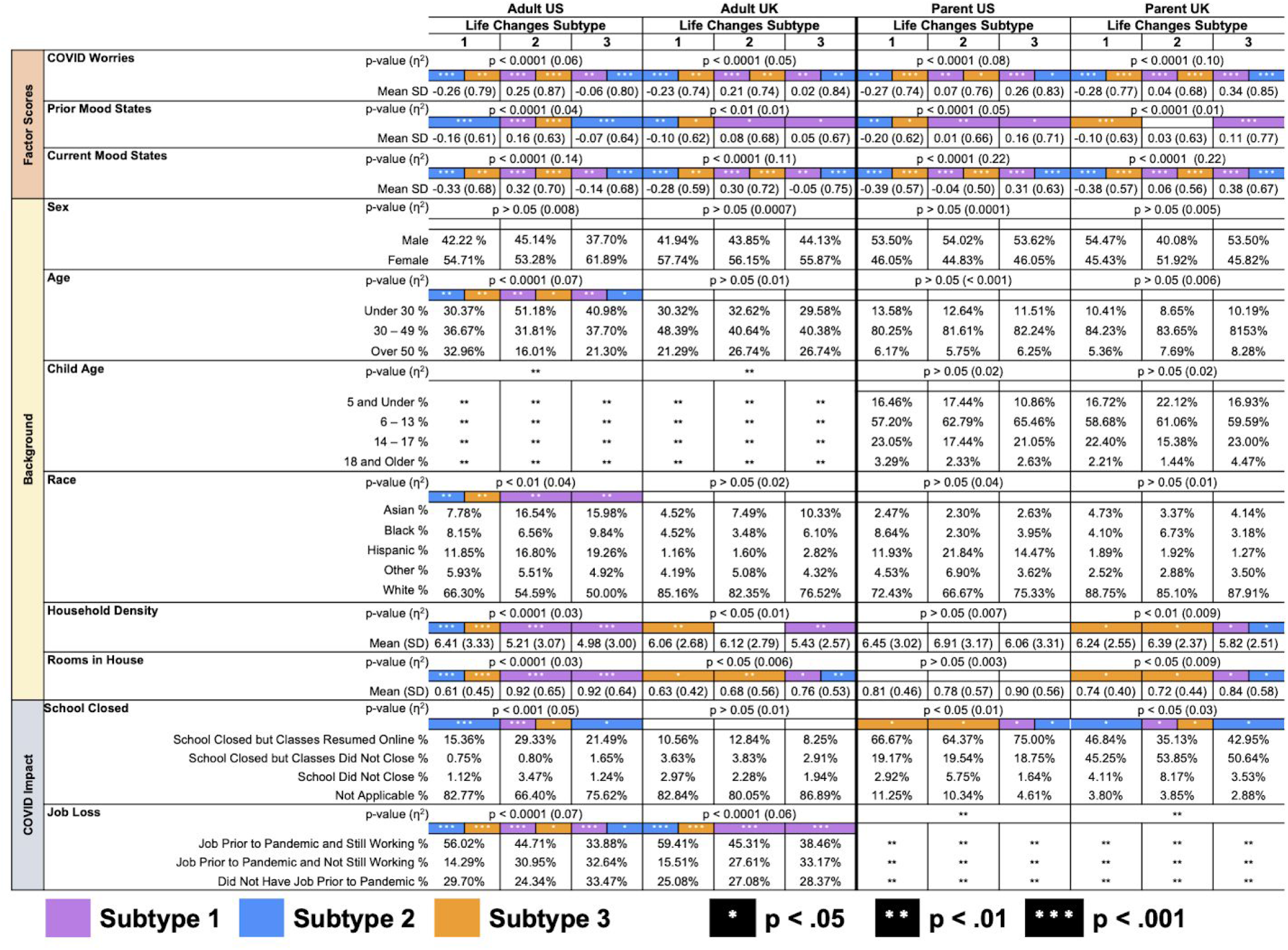
Mean factor scores, demographic characteristics, and pandemic-related school closure and job loss by Life Changes subtypes. Color indicates group in column is significantly different from subtype corresponding to color indicated. Pairwise group differences are represented by white asterisks: * *p <*.05, ** *p* <.01, \*\*\**p* <.001.

### 3.5 Predicting Mood States in the Early Phase of the Pandemic (Aim 4)

The random forests estimation of the relative importance of demographics, Prior Mood States, COVID Worries, Life Changes subtype, and Prior Behavior, Media, and Substance Use, in predicting Current Mood States are shown in Figure 2. Variables are ranked along the y-axis according to their importance in predicting out-of-bag Current Mood States as measured by the percent change in MSE, displayed on the x-axis. Models accounted for a high percentage of the out-of-bag variance in Current Mood States across all samples (Adult; US: 42.2%, UK: 48.7%; Parent; US: 52.6%, UK: 44.6%).

**Figure 2:**
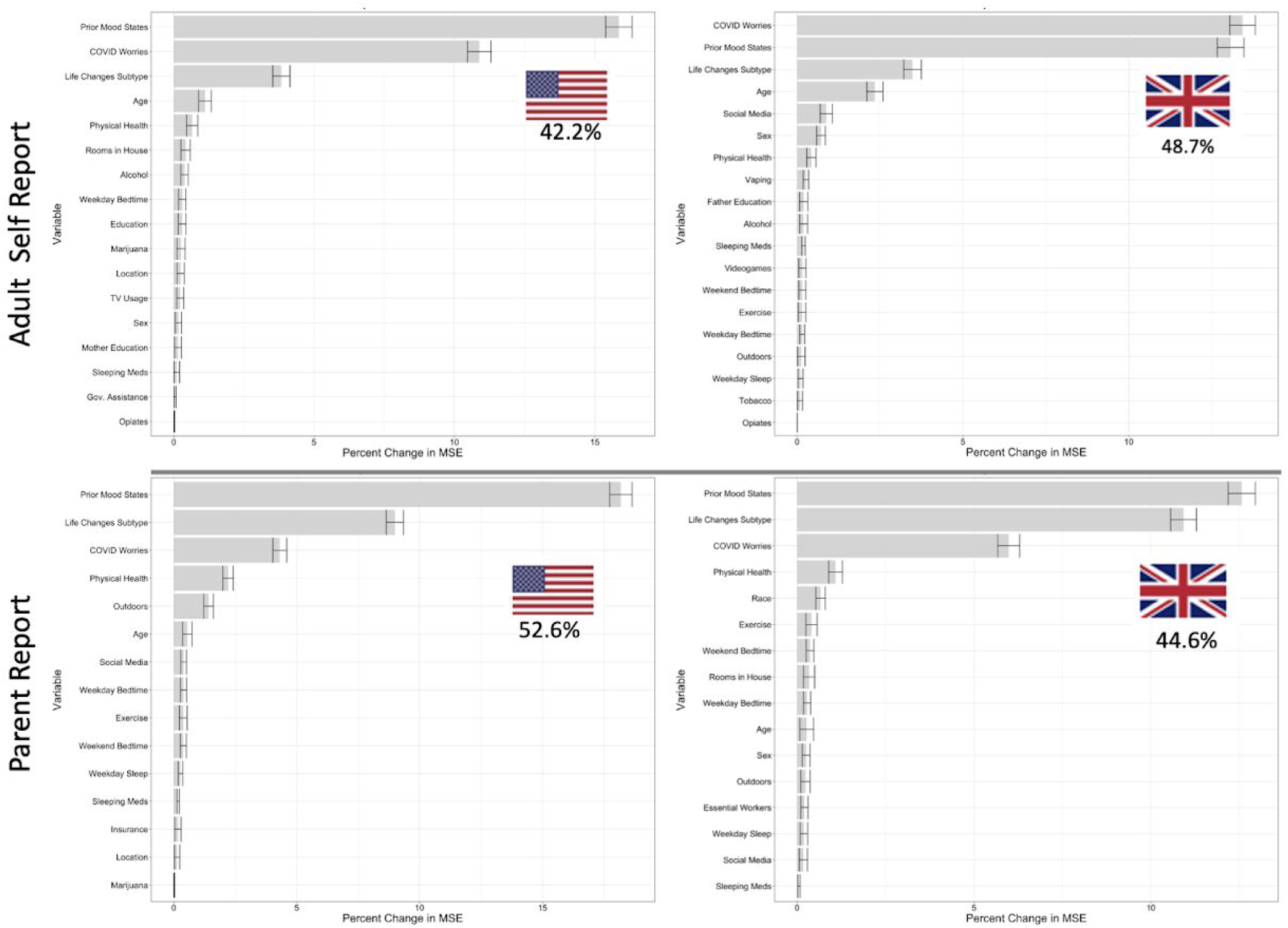
Variable importance and overall performance of Random Forest models predicting Current Mood States in the US and UK in both Adult self-report and parent report data. Variables are ranked by importance as measured by out-of-bag change in mean squared error (MSE), and those with a 95% lower bound above zero are shown here. Variables included: Prior Mood States, Life Changes Subtype, COVID Worries, physical health, age, sex, outdoors, exercise, social media, TV, videogame, weekend bedtime, weekend sleep, weekday bedtime, weekday sleep, insurance, rooms in house, government assistance, number in household, essential worker in household, Marijuana, Alcohol, vaping, opiates, sleeping medication and other drug use.

Prior Mood States, COVID Worries, Life Changes Subtype, and age were the most important domains for predicting Current Mood States in descending order of importance in the adult sample. On the other hand in the parent report sample, after prior Mood states, the Life Changes subtype, COVID Worries and parent-rated health predicted Mood States. Across the US and UK these importance values were highly replicable (Adult Self Report: MSE Pearson’s r = 0.98; Impurity Pearson’s r = 0.99. Parent Report: MSE Pearson’s r = 0.96; Impurity Pearson’s r = 0.98).

To assess the successive impact of adding COVID Worries and the Life Changes Subtype to our predictions, we tested a baseline model without these variables and found much lower predictive accuracy (Adult; US: 30.4%, UK: 28.0%; Parent; US: 35.8%, UK: 20.7%). Performance increased dramatically when adding either COVID Worries (Adult; US: +10.0%, UK: +16.8%; Parent; US: +7.9%, UK: +12.4%) or Lifestyle Changes Subtype (Adult; US: +5.5%, UK: +7.2%; Parent; US: +13.3%, UK: +17.4%). As expected based on the performance and variable importance ranking of the full models, COVID Worries conferred more additional predictive performance for the Adult sample, while Lifestyle Changes Subtype conferred more additional predictive performance for the Parent report sample.

Based on the random forest results, the 4 most important predictors of Current Mood States in adults were Prior Mood States, COVID Worries, Life Changes Subtype, and age; in children, next most important was a variable of parent-rated health of the child. Our trained linear model, including interactions, was able to predict between 49.6-56.5% of the variance in Current Mood States across all four of the ⅓ sample hold-out data. This result strongly demonstrates the generalizability of the importance of these variables in predicting Current Mood States.

## DISCUSSION

The findings presented here demonstrate the feasibility, reliability, and construct validity of the CRISIS in large pilot samples in the US and UK. The high completion rates, low rates of missing data, and rapid completion times demonstrate that the CRISIS is feasible to administer in large samples. The unidimensional Mood States and COVID Worries factor scores reached excellent levels of both internal and test retest reliability (Omega > 0.9; ICCs between 0.79-0.87), and individual items from other measured domains showed high ICC as well. High reliability of survey instruments is absolutely critical in evaluating change and in robustly identifying those in need of interventions or other forms of support. The unidimensional structure of the COVID Worries and Mood States domains were highly replicable across all samples. The construct validity of the CRISIS was demonstrated by the reproducible associations between measured domains as well as the associations of COVID Worries and pandemic-associated life changes with Current Mood States determined from the well-established circumplex model of affect.^27,28^ Together, the results demonstrate the utility of the CRISIS for population-based mental health research during the COVID-19 pandemic. The highly robust replication of our findings across samples, countries, and informants suggests that CRISIS would be appropriate for application across an array of research settings around the world. To date, the CRISIS is being administered in more than eight countries and translations have been developed in several languages. Thus, we will have the opportunity to test the reliability and validity in middle- and low-income countries around the globe. More about CRISIS and its adaptation for Autism and related Neurodevelopmental Conditions (CRISIS AFAR; www.crisissurvey.org/), and other such international collaborations through the Wellcome Trust (www.COVIDminds.org/) can be found online.

COVID Worries was either the first (UK) or second (US) most important predictor of Current Mood States among adults in April, 2020, followed by pandemic-associated life changes. These results suggest that fear and worry about COVID and resulting changes in routines and daily life are significant drivers of adverse mental health outcomes associated with the pandemic, consistent with previous data on the impact of the Fukushima disaster.^37,38^ This speaks to the value of measuring COVID-related fears and worries, as in the CRISIS and other instruments developed for the COVID pandemic.^18-24^ It also implies that active steps that could be taken to offset the impact and lessen the burden of changes in lifestyle by social, governmental or other agencies could have a significant impact in ameliorating negative mental health outcomes.^39^ Future studies including repeated longitudinal assessments could assess the potential long-term effects of such policies on mental health and enable comprehensive evaluation of costs and benefits.

Our finding of parent report data that indicated that Current Mood States among children was more strongly related to Life Changes than COVID Worries is consistent with the known importance of regular, predictable, daily routines for pediatric mental health,^40,41^ and suggests that attending to changes in children’s lives may be key to predicting those at greatest risk for negative psychological impact of the pandemic. Consistent with the review of Brooks et al,^7^ subgroups reporting family and social isolation stress in both adults and children in the US and UK had significantly higher Current Mood States scores. In addition, subgroups of children with higher family and social isolation stress also experienced the highest parent-reported stress related to financial and food security. This underscores the impact of multifactorial physical, emotional, interpersonal, social, and financial stressors that converge during this pandemic. The links between Life Changes profiles and the COVID Worries and Mood States factors attest to the validity of these domains and their potential utility as targets for pandemic-related interventions.

Possessing information on mental and behavioral health prior to the pandemic significantly enhances the ability to assess the impact of the pandemic and its correlates on mental health outcomes by allowing researchers to evaluate both the change in mental health during the pandemic and the potential for prior characteristics to moderate the effects of pandemic-related stressors. Indeed, this is a central goal of the CRISIS initiative. Because information on prior mental health may not be available to all researchers, we included retrospective reports of key domains in the CRISIS to enable researchers to evaluate the role of prior characteristics and clinical state.^42^ Our finding that Prior Mood States and Prior Habits were significantly associated with Current Mood States provides support for the importance of psychological status prior to the pandemic. The reproducibility of the structure of the multi-dimensional domains of Life Changes and Prior Habits attests to the value of this feature of the CRISIS. Ultimately, prospective measures of pre-COVID mental health will facilitate the identification of those at greatest risk of long term sequelae of this pandemic, as shown in previous disaster research.^26^

This study is limited by its use of a web-based convenience sample, which raises the possibility of selection bias and impedes our ability to generalize our findings to the broader US and UK populations. This limitation applies to the majority of the current mental health surveys of COVID-19, which have mostly used samples ascertained through web-based sources.^43^ We employed this approach in order to quickly deploy the CRISIS to large numbers of participants within a brief time frame during the initial peak of the pandemic, to rapidly evaluate test-retest reliability, and to pilot the shorter follow-up version of the CRISIS. We do not expect the composition of our sample to strongly affect our findings regarding the structure of the CRISIS, the reliability of its items or unidimensional domains, or its construct validity. The relatively lower racial/ethnic diversity in the present study is an important limitation, particularly in light of the disproportionate impact of COVID-19 on marginalized communities.^44,45^ Further work is required to assess the properties of CRISIS in different racial/ethnic groups, cultural settings, and languages. However, these data source provided samples with broad coverage of the US and UK populations with respect to age, sex, and race.^44,46,47^ Moreover, this pilot study did not include the youth self-report version of the CRISIS, but this work is now underway in our collaborative network. Validation of youth reports will be particularly relevant because the profiles of predictors of change derived from families with children under age 18 differed from those from adult households.

These findings reflect the initial steps of instrument development to implement our collaborative effort on the mental health impact of the COVID-19 pandemic. With some exceptions,^18-24^ this study of the CRISIS is one of the few COVID-19 stress/anxiety questionnaires that has provided psychometric data on its factor structure and/or validity of the content. The factors derived in our study replicate those of Taylor *et al* ^21^ who likewise employed statistical approaches that demonstrated the heterogeneity of impact of COVID-19 fears and anxiety, and the impact of prior mental health problems on its severity. We further demonstrate the utility of the CRISIS through its reproducible structure, its acceptability, feasibility, and reliability, and its construct validity across multiple samples. The inclusion of adult and parent versions to examine differences in the impact of COVID-19 across the life span will also facilitate our ability to gain insight into the impact of the pandemic on children and families. Efforts to administer the survey in previously well characterized samples such as the Healthy Brain Network, a landmark ongoing mental health study of 10,000 children with deep phenotyping across a range of psychiatric, cognitive, affective, language, genetic, and neuroscientific characteristics,^48^ are underway. The major goal of our initiative is to conduct research that informs priorities for interventions and policy changes to ameliorate the mental health consequences of the pandemic, both acutely and in the long term.

## Data Availability

Data will be made publicly available upon request.

## Acknowledgements

We would like to thank the families that participated in the CRISIS Survey. *Disclaimer:* The views and opinions expressed in this article are those of the authors and should not be construed to represent the views of any of the sponsoring organizations, agencies, or the US government.

## Supplemental Materials

### Supplemental Methods

#### Overview

First, we used confirmatory factor analysis to assess the structure of each of the domains and inform the calculation of factor scores for unidimensional constructs. Test-retest reliability was calculated for unidimensional factors and individual items using ICC(3,1).^30^ We used Louvain community detection, a clustering technique, to meaningfully summarize domains for which unidimensional factors exhibited poor fit. Finally, we used random forests to demonstrate the construct validity of the CRISIS by assessing the importance of the included domains in predicting the Current Mood States factor.

#### Subtyping

We use bagging-enhanced Louvain Community Detection to discover groups of individuals that have profiles across both the Life Changes questions, and the Daily Behaviors, Media Use, and Substance Use questions, which we called the Prior Habits subtypes. Louvain Community Detection is known to robustly link observations together through the use of an iterative modularity-optimizing procedure to find groups of individuals. Other clustering approaches, such as K-means, or spectral clustering, require the experimenter to choose the resolution of the clustering a priori, which can be problematic and lead to instability across samples.^49-51^ Louvain Community detection on the other hand, through iterative permutations of individuals in each community, optimizes for modularity, a commonly used metric of cluster quality.^32,52^ We enhance the reproducibility of our subtyping method through the use of bootstrap aggregation, or bagging. Using bootstrap aggregated clustering creates more reproducible clusters by reducing variability that may occur due to random variations in sample composition.

**S. Figure 1:**
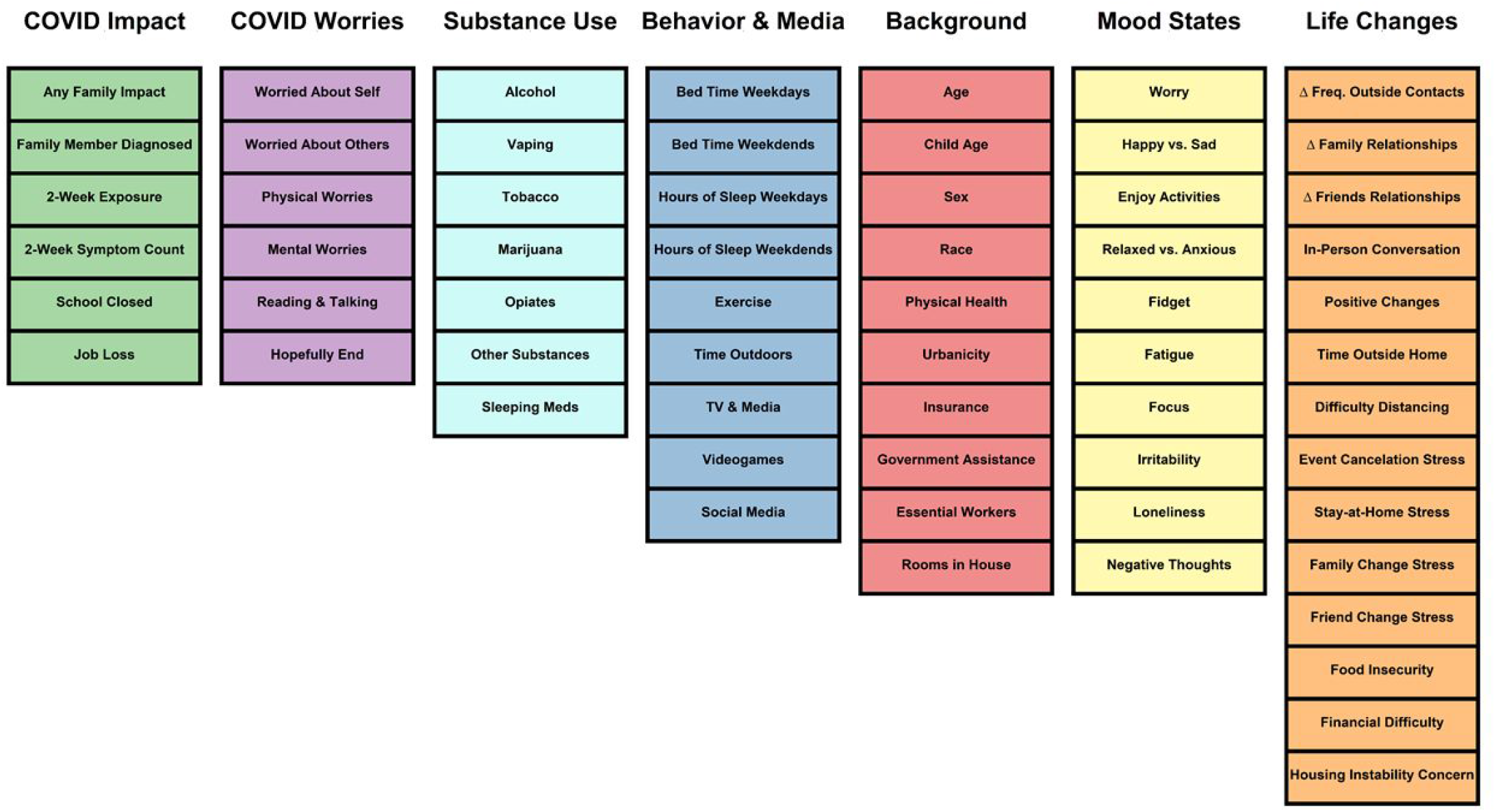
Columns contain the variables included in each of the individual categories and colored by category respectively.

#### Random Forest

Briefly, the RF algorithm creates a series of decision trees for which a random selection of variables are chosen and a bootstrapped sample is used to train the model. For each iteration of the 1000 bootstrap runs, the performance on each of these decision trees on the out-of-sample data, roughly ⅓ of the sample, is aggregated and used to assess the performance of the RF model. (For a review of RF, see^51^). RF provides a robust assessment of the relative impact of each of these variables in predicting outcomes, known as variable importance, which we assess for each variable in our predictive model. To protect against overfitting, we create a null performance distribution from our own data by shuffling the outcome variable and repeating the random forest prediction pipeline 1000 times. The out-of-sample prediction R-squared value is then calculated for our prediction and compared to the distribution of these 1000 shuffled null models. We assess if our predictive accuracy surpasses the 99.9999% confidence interval of the null model. We used COVID Worries and Prior Mood States transformed into quintiles in order to protect against the inflation effects that RF can have on the variable importance of continuous versus categorical variables.^53^

### Supplemental Results

#### Prior Habits Subtyping

We found 3 Prior Habits subtypes in both US and UK adult samples. Individuals in subtype 1 spend the least amount of time exercising and outdoors, and in the US go to sleep relatively later and spend more time on media, while in the UK this subtype goes to sleep earlier and spends an average amount of time on media. Subtype two goes to bed later, in the US this subtype gets relatively less sleep but spends more time exercising and outdoors, while in the UK this group gets an average amount of sleep and time spent exercising and outdoors. Subtype 3 goes to bed earliest and spends more time exercising and outdoors, and reports relatively lower media use and drug use.

Across US and UK samples these subtypes were highly reproducible, with high Pearson’s correlations between subtype mean scores (r 0.71 – 0.96) for the adult sample. ANOVA revealed Prior Habits subtypes had different Prior Mood States factor (US p< 0.00001, UK p< 0.00001) but ANCOVA of Current Mood States controlling for Prior Mood states only differed in the US sample (US p > 0.05, UK p > 0.05). The US sample also showed different Mood States scores by subtype (US p < 0.00001; UK p = 0.06). COVID Worries scores differed by subtype in the UK but not US (US p > 0.05, UK p < 0.01). Mean factor scores by subtype are shown in the Supplemental table 3. Overall, results indicated significant differences between subtypes in prior mood states scores in both US and UK, with subtype 1 showing highest scores in the US and subtype 3 showing highest scores in the UK. We also see significant subtype differences by age in both US and UK. Prior Habits subtypes also differed by key demographic variables including age, race, education, rooms in house, household density, and employment status (see Supplement).

The parent report US and UK subtypes were highly reproducible, with high Pearson’s correlations between subtype mean scores (r 0.97 – 0.99) In the parent report data we found 3 Prior Habit subtypes in the US sample and 4 in the UK sample. Individuals in subtype 1, in both the US and UK went to bed later in the evening and got the least amount of sleep, and also spent less time exercising and outdoors. Subtype 1 also reports the highest media use and ratings of drug use. Subtype 2 went to bed early and got above average sleep, but below average exercise, outdoor time, and social media use. Compared to previous subtypes, Subtype 3 showed relatively divergent patterns across the US and UK, with the US sample showing later than average bedtime and media use, less than average sleep, while in the UK individuals had an early bedtime but greater than average sleep and less media use on average. Subtype 4, in the UK only, showed the earliest bedtime, and the greatest amount of sleep, exercise and outdoor time, and the least amount of media use.

One-way ANOVA of subtype by Prior Mood score and ANCOVA of Current Mood factor score, controlling for Prior Mood Score, indicates that these Prior Habits subtypes show different patterns of Mood States over time (Prior Mood State: US p > 0.05, UK p< 0.00001; Current Mood State: US p > 0.05, UK p > 0.05). One-way ANOVA of subtype by COVID Worries factor score shows that these Prior Habits subtypes are sensitive to differences in COVID worries in the UK but not US (US p < 0.05, UK p < 0.00001). The parent report subtypes show subtype 1 and 2 with the highest and lowest COVID worries factor scores respectively. We also see that subtype 1 and 2 are significantly different in age distribution, with subtype 1 having the most teenagers and subtype 2 having the most children under 5.

**S Figure 2.**
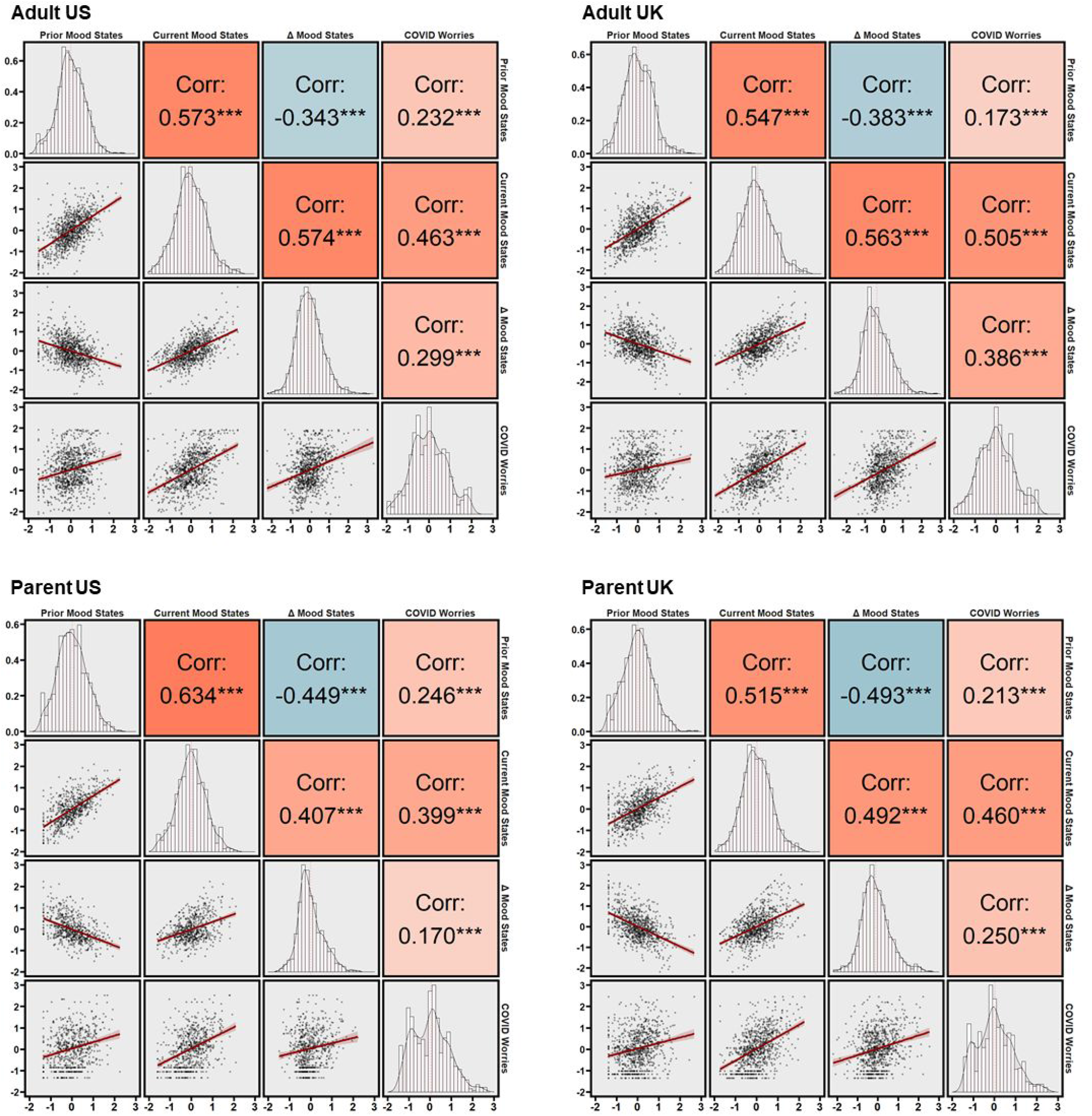
Correlations between all factors scores (Prior Mood States, Current Mood States, Δ Mood States, and COVID Worries) across all samples. Lower left panels display matrices of scatter plots of the correlation between the factors. X and Y label values represent a standard loading of −2 at the origin and 3 at the maximum for each of the factors. Corresponding diagonal panels show the histogram distribution of the factor correlations. Pearson correlation coefficient values are presented in the top right panels.

**S. Table 1.** We summarize missing data here. First, the number of items missing on average in each completed survey; Second, the range of missing items across all completed surveys, and third the average time to completion of the surveys (in minutes). Fourth, the total number of incomplete surveys for each sample.

|  | US Adult | UK Adult | US Parent | UK Parent |
| --- | --- | --- | --- | --- |
| **Missing Data** |  |  |  |  |
| Mean (SD) | 0.6 (1.8) | 0.5 (1.1) | 0.4 (0.9) | 0.4 (1.0) |
| **Range of Missed Items** | 0-54 | 0-17 | 0-14 | 0-12 |
| **Time to Completion** |  |  |  |  |
| Mean (SD) | 13.9 (13.1) | 14.4 (11.5) | 14.1 (7.5) | 13.9 (20.9) |
| **Incomplete Observations** |  |  |  |  |
| N (%) | 108 (6.0) | 43 (2.6) | 43 (3.5) | 88 (5.4) |

**S Table 2:**
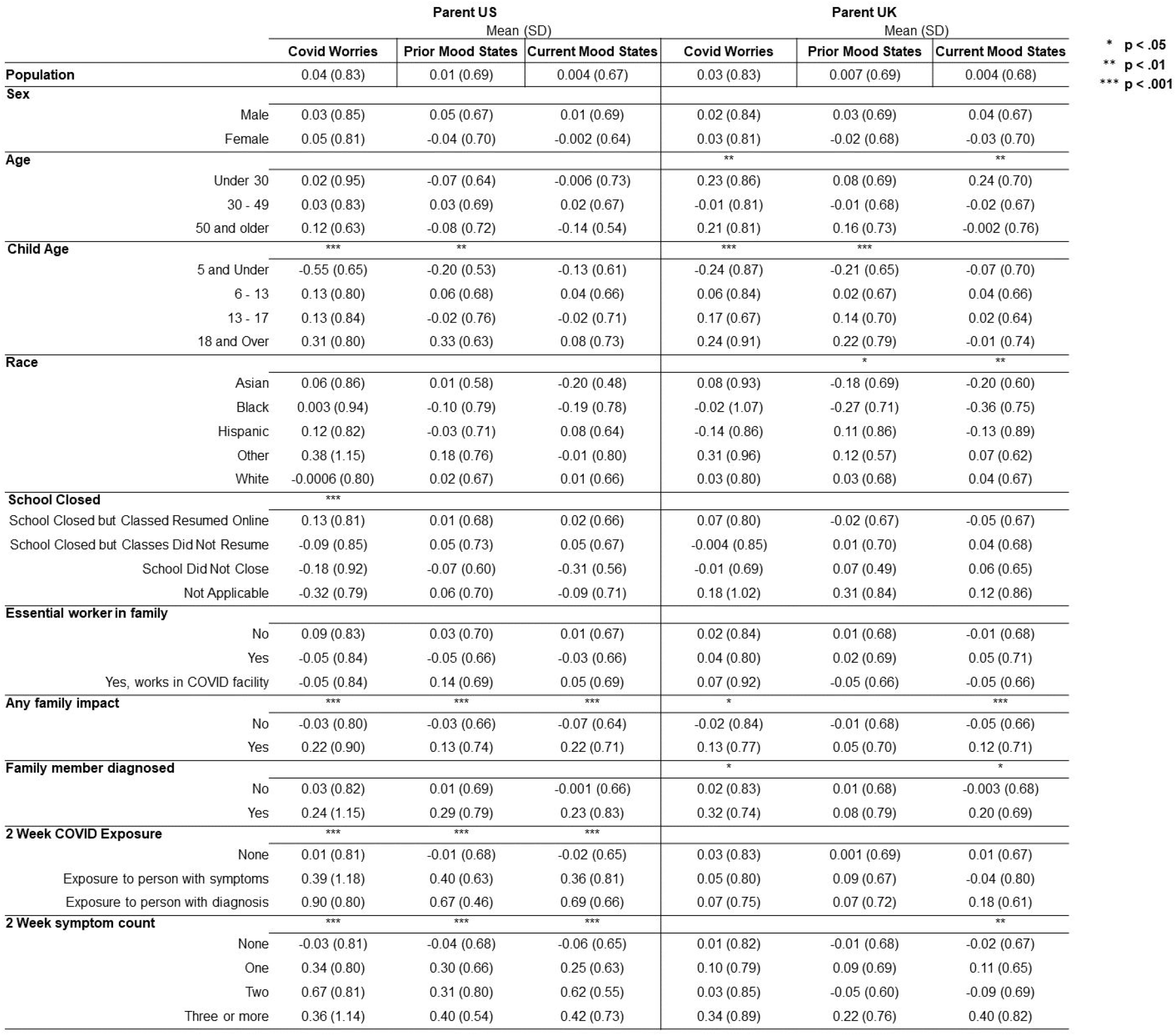
Parent report overall mean and SD of factor scores (COVID Worries and Mood States) followed by mean and SD by demographic group and COVID-related characteristics. Significant ANOVA demographic group differences are represented by asterisks; * *p <*.05, ** *p* <.01, \*\*\**p* <.001.

**S Table 3.**
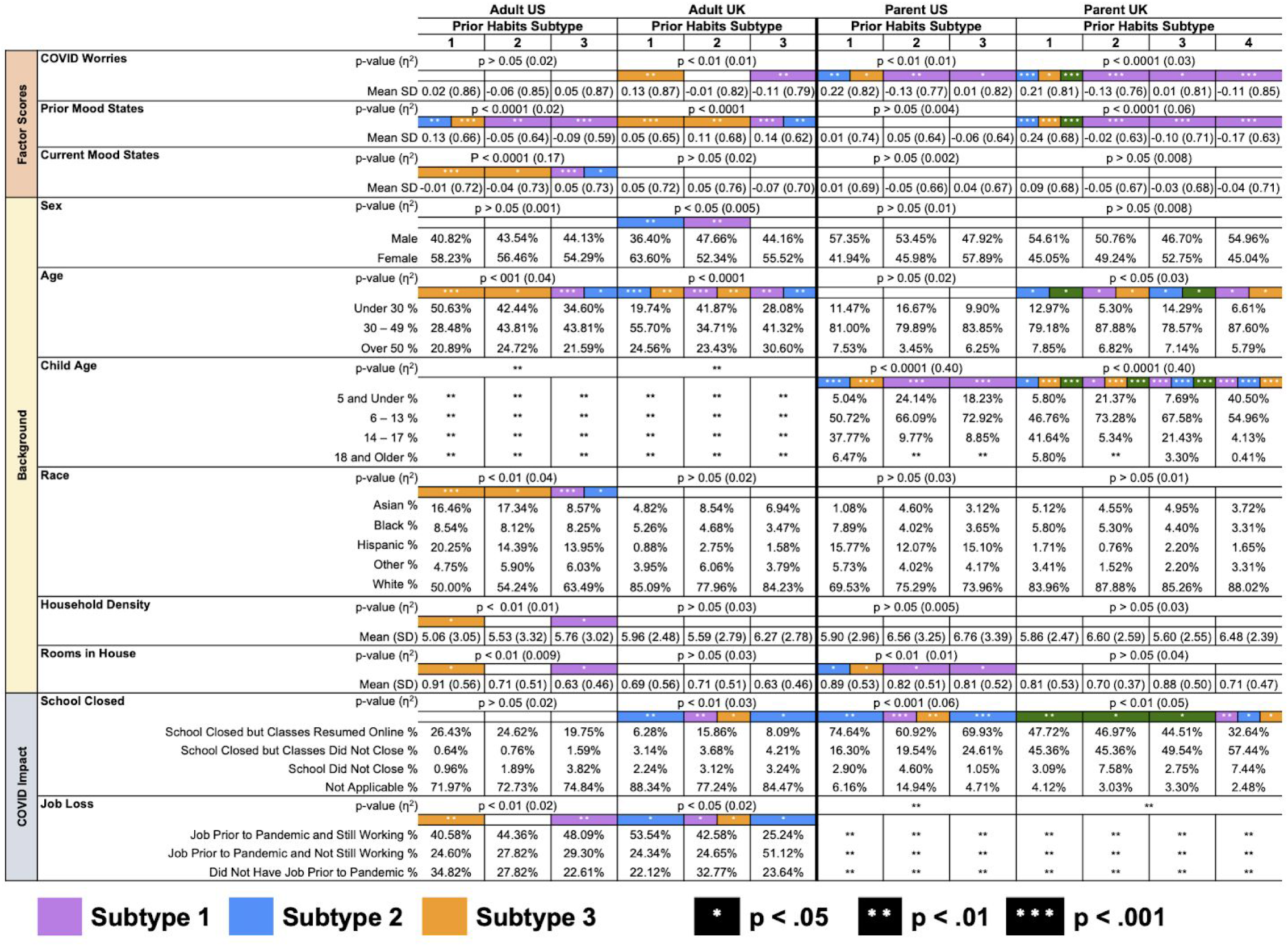
Prior Habits subtypes are indicated by color (Subtype 1, purple; Subtype 2, blue; Subtype 3, orange). Significant ANOVA group differences (COVID Worries, Prior Mood States, and Current Mood States) and Chi-Square group differences (Sex, Age, Child Age, Race, School Closed, and Job Loss) are represented by white asterisks; * *p <*.05, ** *p* <.01, ***p <.001 and by color according which subtype significant differences were observed.

**S Table 4.** Intraclass Correlation Coefficient (ICC) mean and standard deviation for Behavior & Media, Life Changes, and Substance Use variables.

| Life Changes ICC |  | Behavior & Media ICC |  |  | Substance Use ICC |  |  |
| --- | --- | --- | --- | --- | --- | --- | --- |
|  | Current<br>Mean (SD) |  | Prior<br>Mean (SD) | Current<br>Mean (SD) |  | Prior<br>Mean (SD) | Current<br>Mean (SD) |
| Positive Change | 0.66 (0.07) | Bedtime Weekdays | 0.78 (0.06) | 0.86 (0.08) | Alcohol | 0.83 (0.19) | 0.66 (0.32) |
| Time Outside | 0.66 (0.08) | Bedtime Weekends | 0.80 (0.07) | 0.86 (0.04) | Vaping | 0.92 (0.11) | 0.98 (0.02) |
| Contacts Changed | 0.36 (0.12) | Hours of Sleep Weekdays | 0.80 (0.05) | 0.79 (0.08) | Tobacco | 0.99 (0.01) | 0.98 (0.07) |
| Family Change | 0.58 (0.06) | Hours of Sleep Weekends | 0.78 (0.04) | 0.77 (0.09) | Marijuana | 0.98 (0.03) | 0.95 (0.05) |
| Friends Change | 0.70 (0.13) | Exercise | 0.76 (0.05) | 0.82 (0.07) | Opiates | 0.80 (0.36) | 0.78 (0.37) |
| Cancelations Difficulty | 0.64 (0.12) | Time Outdoors | 0.75 (0.04) | 0.82 (0.06) | Other Substances | 0.95 (0.05) | 0.92 (0.09) |
| Living Difficulty | 0.65 (0.12) | TV & Media | 0.62 (0.14) | 0.63 (0.12) | Sleeping Meds | 0.86 (0.16) | 0.82 (0.12) |
| In-Person Conversation | 0.84 (0.21) | Social Media | 0.82 (0.05) | 0.85 (0.04) |  |  |  |
| Restriction Stress | 0.68 (0.08) | Video Game | 0.84 (0.04) | 0.86 (0.03) |  |  |  |
| Distancing Difficulty | 0.48 (0.13) |  |  |  |  |  |  |
| Family Change Stress | 0.60 (0.09) |  |  |  |  |  |  |
| Friends Change Stress | 0.62 (0.08) |  |  |  |  |  |  |
| Financial Difficulty | 0.78 (0.06) |  |  |  |  |  |  |

**S. Figure 3:**
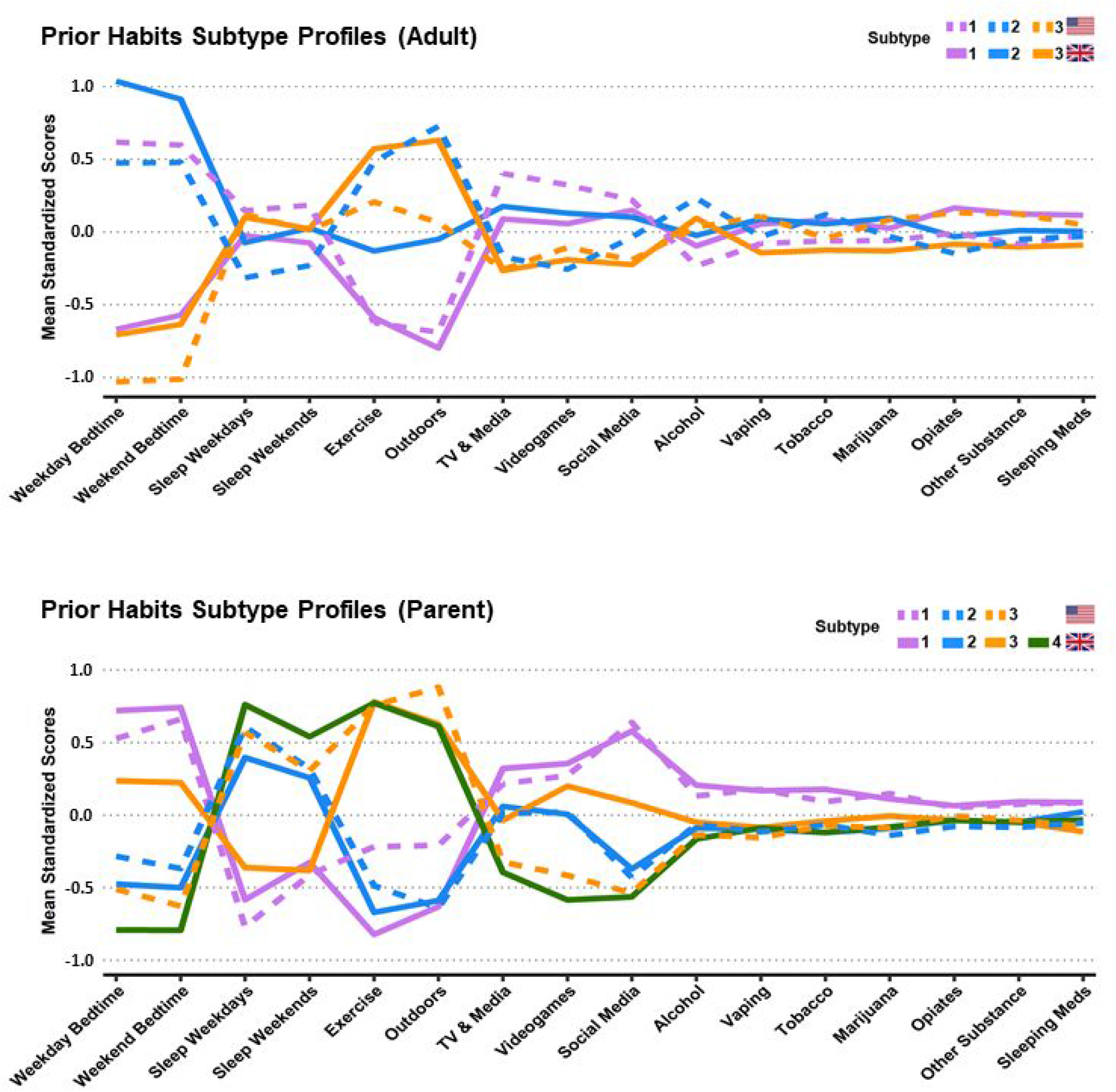
Prior Habit Subtype profiles from adult self-reports and parent reports. Mean normalized profile loadings are displayed on the y-axis. US subtypes in solid lines, UK in dashed lines. Notes: Δ Family Relations and Δ Friends Relationships were reverse coded to facilitate Subtype interpretation; higher scores indicate worsening quality of the relationships. Prior to the community detection analyses In-Person Conversation was re-coded into tertiles.

**S. Table 5.** Random Forest tests identified the above five variables (COVID Worries, Prior Mood States, Life Changes Subtypes, Age, and Physical Health) to be the most important for predicting Current Mood States. The relationship between COVID Worries, Prior Mood States and Current Mood States was tested with Pearson correlations in and out of sample. ANOVA was used to test the difference in Current Mood States with Age (Adult Report) and parent-rated Physical Health (Parent Report) in out of sample. P values <.05 indicate significant differences in membership between Prior Habits and Life Changes subtypes. Correlation values are provided next the p value for COVID Worries and Prior Mood States.

|  | US |  | UK |  |
| --- | --- | --- | --- | --- |
|  | In Sample | Out of Sample | In Sample | Out of Sample |
| **Adult Report** |  |  |  |  |
| **COVID Worries (Correlation)** | p < .00001, 0.46 | p < .00001, 0.46 | p < .00001, 0.51 | p < .00001, 0.50 |
| **Prior Mood States (Correlation)** | p < .00001, 0.57 | p < .00001, 0.58 | p < .00001, 0.55 | p < .00001, 0.54 |
| **Lifestyle Changes Subtypes** | p < .00001 | p < .00001 | p < .00001 | p < .00001 |
| **Age** | p < .00001 | p < .00001 | p < .00001 | p < .00001 |
| **Parent Report** |  |  |  |  |
| **COVID Worries (Correlation)** | p < .00001, 0.40 | p < .0005, 0.44 | p < .00001, 0.46 | p < .00001, 0.47 |
| **Prior Mood States (Correlation)** | p < .00001, 0.64 | p < .00001, 0.61 | p < .00001, 0.51 | p < .00001, 0.53 |
| **Lifestyle Changes Subtypes** | p < .00001 | p < .00001 | p < .00001 | p < .00001 |
| **Physical Health** | p < .00001 | p < .00001 | p < .00001 | p < .00001 |

**S. Table 6.** Chi-Square tests were conducted between the Prior Habits and Life Changes subtypes to assess the existence of a differential makeup. P values <.05 indicate significant differences in membership between Prior Habits and Life Changes subtypes.

|  | Adult US | Adult UK | Parent US | Parent UK |
| --- | --- | --- | --- | --- |
| **Subtype Homogeneity** | p < .00001 | p < .00001 | p < .00001 | p < .005 |

## BIBLIOGRAPHY

1. Peeri, N. C. et al. The SARS, MERS and novel coronavirus (COVID-19) epidemics, the newest and biggest global health threats: what lessons have we learned? Int. J. Epidemiol. 49, 717–726 (2020).

2. Goldmann, E. & Galea, S. Mental Health Consequences of Disasters. Annu. Rev. Public Health 35, 169–183 (2014).

3. Norris, F. H. et al. 60,000 Disaster Victims Speak: Part I. An Empirical Review of the Empirical Literature, 1981-2001. Psychiatry: Interpersonal and Biological Processes 65, 207–239 (2002).

4. North, C. S. & Pfefferbaum, B. Mental Health Response to Community Disasters: A Systematic Review. JAMA 310, 507 (2013).

5. Neria, Y., Nandi, A. & Galea, S. Post-traumatic stress disorder following disasters: a systematic review. Psychol. Med. 38, 467–480 (2008).

6. Bromet, E. J. et al. Post-traumatic stress disorder associated with natural and human-made disasters in the World Mental Health Surveys. Psychol. Med. 47, 227–241 (2017).

7. Brooks, S. K. et al. The psychological impact of quarantine and how to reduce it: rapid review of the evidence. Lancet 395, 912–920 (2020).

8. Frasquilho, D. et al. Mental health outcomes in times of economic recession: a systematic literature review. BMC Public Health 16, 115 (2015).

9. Rogers, J. P. et al. Psychiatric and neuropsychiatric presentations associated with severe coronavirus infections: a systematic review and meta-analysis with comparison to the COVID-19 pandemic. The Lancet Psychiatry S2215036620302030 (2020).

10. Liu, C. H., Zhang, E., Wong, G. T. F., Hyun, S. & Hahm, H. ‘chris’. Factors associated with depression, anxiety, and PTSD symptomatology during the COVID-19 pandemic: Clinical implications for U.S. young adult mental health. Psychiatry Res. 290, 113172 (2020).

11. Asmundson, G. J. G. & Taylor, S. How health anxiety influences responses to viral outbreaks like COVID-19: What all decision-makers, health authorities, and health care professionals need to know. J. Anxiety Disord. 71, 102211 (2020).

12. Mazza, M., Marano, G., Lai, C., Janiri, L. & Sani, G. Danger in danger: Interpersonal violence during COVID-19 quarantine. Psychiatry Res. 289, 113046 (2020).

13. Araújo, F. J. de O., de Lima, L. S. A., Cidade, P. I. M., Nobre, C. B. & Neto, M. L. R. Impact Of Sars-Cov-2 And Its Reverberation In Global Higher Education And Mental Health. Psychiatry Res. 288, 112977 (2020).

14. Luo, M., Guo, L., Yu, M. & Wang, H. The Psychological and Mental Impact of Coronavirus Disease 2019 (COVID-19) on Medical Staff and General Public – A Systematic Review and Meta-analysis. Psychiatry Res. 113190 (2020).

15. Troyer, E. A., Kohn, J. N. & Hong, S. Are we facing a crashing wave of neuropsychiatric sequelae of COVID-19? Neuropsychiatric symptoms and potential immunologic mechanisms. Brain Behav. Immun. S088915912030489X (2020).

16. Fancourt, D. Longitudinal studies | COVID-MIND. COVID-MIND https://www.covidminds.org/longitudinal-studies (2020).

17. Living systematic review of mental health in COVID-19. https://www.depressd.ca/covid-19-mental-health (2020).

18. Lee, S. A. Coronavirus Anxiety Scale: A brief mental health screener for COVID-19 related anxiety. Death Stud. 44, 393–401 (2020).

19. Qiu, J. et al. A nationwide survey of psychological distress among Chinese people in the COVID-19 epidemic: implications and policy recommendations. General Psychiatry 33, e100213 (2020).

20. Ahorsu, D. K. et al. The Fear of COVID-19 Scale: Development and Initial Validation. Int. J. Ment. Health Addict. (2020) doi:10.1007/s11469-020-00270-8.

21. Taylor, S. et al. Development and initial validation of the COVID Stress Scales. J. Anxiety Disord. 72, 102232 (2020).

22. Pérez-Fuentes, M. del C. et al. Questionnaire on Perception of Threat from COVID-19. J. Clin. Med. Res. 9, 1196 (2020).

23. Repišti, S. et al. How to measure the impact of the COVID-19 pandemic on quality of life: COV19-QoL – the development, reliability and validity of a new scale. Global Psychiatry -1, (2020).

24. Kira, I. A. et al. Measuring COVID-19 as Traumatic Stress: Initial Psychometrics and Validation. J. Loss Trauma 1–18 (2020).

25. Holmes, E. A. et al. Multidisciplinary research priorities for the COVID-19 pandemic: a call for action for mental health science. The Lancet Psychiatry 0, (2020).

26. Gordon, J. A. & Borja, S. E. The COVID-19 Pandemic: Setting the Mental Health Research Agenda. Biological psychiatry vol. 88 130–131 (2020).

27. Larsen, R. J. & Diener, E. Promises and problems with the circumplex model of emotion. (1992).

28. Posner, J., Russell, J. A. & Peterson, B. S. The circumplex model of affect: An integrative approach to affective neuroscience, cognitive development, and psychopathology. Dev. Psychopathol. 17, 715 (2005).

29. Peer, E., Brandimarte, L., Samat, S. & Acquisti, A. Beyond the Turk: Alternative platforms for crowdsourcing behavioral research. J. Exp. Soc. Psychol. 70, 153–163 (2017).

30. Shrout, P. E. & Fleiss, J. L. Intraclass correlations-uses in assessing rater reliability. Psychol. Bull. 86, 420–428 (1979).

31. Brunner, M., Nagy, G. & Wilhelm, O. A tutorial on hierarchically structured constructs. J. Pers. 80, 796–846 (2012).

32. Blondel, V. D., Guillaume, J.-L., Lambiotte, R. & Lefebvre, E. Fast unfolding of communities in large networks. J. Stat. Mech. 2008, P10008 (2008).

33. Nikolaidis, A. et al. Bagging improves reproducibility of functional parcellation of the human brain. Neuroimage 116678 (2020).

34. Domingos, P. M. Why Does Bagging Work? A Bayesian Account and its Implications. in KDD 155–158 (Citeseer, 1997).

35. Breiman, L. Random Forests. Mach. Learn. 45, 5–32 (2001).

36. Probst, P., Wright, M. N. & Boulesteix, A. Hyperparameters and tuning strategies for random forest. WIREs Data Mining Knowl Discov 9, 281 (2019).

37. Harada, N. et al. Mental health and psychological impacts from the 2011 Great East Japan Earthquake Disaster: a systematic literature review. Disaster and Military Medicine 1, 17 (2015).

38. Bromet, E. J. Mental health consequences of the Chernobyl disaster. J. Radiol. Prot. 32, N71–N75 (2012).

39. Marroquín, B., Vine, V. & Morgan, R. Mental Health During the COVID-19 Pandemic: Effects of Stay-at-Home Policies, Social Distancing Behavior, and Social Resources. Psychiatry Research 113419 (2020) doi: 10.1016/j.psychres.2020.113419.

40. Spagnola, M. & Fiese, B. H. Family Routines and Rituals: A Context for Development in the Lives of Young Children. Infants Young Child. 20, 284 (2007).

41. Organization, W. H. & Others. Mental health and psychosocial considerations during the COVID-19 outbreak, 18 March 2020. https://apps.who.int/iris/bitstream/handle/10665/331490/WHO-2019-nCoV-MentalHealth-2020.1-eng.pdf (2020).

42. Vindegaard, N. & Benros, M. E. COVID-19 pandemic and mental health consequences: Systematic review of the current evidence. Brain Behav. Immun. (2020) doi:10.1016/j.bbi.2020.05.048.

43. Pierce, M. et al. Says who? The significance of sampling in mental health surveys during COVID-19. The Lancet Psychiatry S2215036620302376 (2020).

44. Williams, D. R. & Cooper, L. A. COVID-19 and Health Equity—A New Kind of ‘Herd Immunity’. JAMA 323, 2478–2480 (2020).

45. Bowleg, L. We’re Not All in This Together: On COVID-19, Intersectionality, and Structural Inequality. Am. J. Public Health 110, 917 (2020).

46. Selden, T. M. & Berdahl, T. A. COVID-19 And Racial/Ethnic Disparities In Health Risk, Employment, And Household Composition: Study examines potential explanations for racial-ethnic disparities in COVID-19 hospitalizations and mortality. Health Aff. 10–1377 (2020).

47. Lassale, C., Gaye, B., Hamer, M., Gale, C. R. & Batty, G. D. Ethnic Disparities in Hospitalization for COVID-19: a Community-Based Cohort Study in the UK. *medRxiv* (2020) doi:10.1101/2020.05.19.20106344.

48. Alexander, L. M. et al. An open resource for transdiagnostic research in pediatric mental health and learning disorders. Sci Data 4, 170181 (2017).

49. Tumer, K. & Ghosh, J. Linear and order statistics combiners for pattern classification. CoRR. *arXiv preprint cs. NE/9905012* (1999).

50. Li, H. et al. K-Means Clustering with Bagging and MapReduce. in 2011 44th Hawaii International Conference on System Sciences 1–8 (ieeexplore.ieee.org, 2011).

51. Friedman, J., Hastie, T. & Tibshirani, R. The elements of statistical learning. vol. 1 (Springer series in statistics New York, 2001).

52. Brandes, U. et al. On Modularity Clustering. IEEE Trans. Knowl. Data Eng. 20, 172–188 (2008).

53. Strobl, C., Boulesteix, A.-L., Zeileis, A. & Hothorn, T. Bias in random forest variable importance measures: illustrations, sources and a solution. BMC Bioinformatics 8, 25 (2007).

